## Supplementary Data for "Metabolic profiling of rheumatoid arthritis neutrophils reveals altered energy metabolism that is not affected by JAK inhibition"

### SUPPLEMENTARY METHODS

#### Chemotaxis assay

The chemotaxis assay was carried out in 24-well tissue culture plates (coated with 12mg/mL polyhema to prevent cell adhesion) using hanging chamber inserts with a 3µM porous membrane, as previously described [18]. fMLP ($10^{-8}$M) was added to RPMI media in the bottom chamber. Neutrophils ($10^6$/mL) were incubated with or without tofacitinib, baricitinib or pan-JAK inhibitor I (all 200 ng/mL) for 30 min prior to addition to the top chamber and incubated for 90 min at 37°C and 5% $CO_2$. The number of migrated cells after 90 min incubation was measured using a Coulter Counter Multisizer-3 (Beckman Coulter).

#### Phagocytosis assay

Neutrophils ($10^6$/mL) were incubated with or without tofacitinib, baricitinib or pan-JAK inhibitor I (all 200 ng/mL) for 30 min prior to the addition of FITC-labelled latex beads (1.1µm diameter) for 30 min (100:1 ratio beads:cells). Cells were centrifuged at 1000g for 3 mins and resuspended in HBSS prior to analysis on a Beckman Coulter CytoFLEX flow cytometer. 10,000 events were analysed.

### SUPPLEMENTARY DATA

**Supplementary Table 1. List of annotated metabolites from neutrophil spectra.** Table reports the total number of annotated peaks for each metabolite and the level of identification based on the metabolite reporting standards set out by the metabolomics society. A representative bin was chosen out of all related annotated peaks and brought forwards in the subsequent statistical analysis. HMDB = Human Metabolome Database

| Metabolite | HMDB ID | Number of peaks annotated in spectrum | MSI level of identification | Representative bin |
|---|---|---|---|---|
| 2 Hydroxyvaleric acid | HMDB0001863 | 12 | 2 | 1.3539 - 1.433 |
| 2-Hydoxybutyric acid | HMDB0000008 | 3 | 2 | 0.9106 - 0.9058 |
| 2-Hydroxy-3-methylpentaonic acid | HMDB0000317 | 24 | 2 | 0.8962 - 0.8504 |
| 3.Hydoxybutyric acid | HMDB0000011 | 2 | 2 | 1.2241 - 1.2168 |
| 3-Hydroxyisovaleric.acid | HMDB0000754 | 2 | 2 | 1.2572 - 1.2498 |
| Acetamide | HMDB0031645 | 1 | 2 | 2.0094 - 2.0039 |
| Acetoatic acid | HMDB0000060 | 1 | 2 | 2.2674 - 2.2618 |
| Acetone | HMDB0001659 | 1 | 2 | 2.2385 - 2.2342 |
| Adipic Acid | HMDB0000448 | 7 | 2 | 1.5445 - 1.5401 |
| ADP | HMDB0001341 | 6 | 1 | 8.5451 - 8.5407 |
| ATP | HMDB0000538 | 9 | 1 | 8.2742 - 8.2719 |
| Choline | HMDB0000097 | 1 | 2 | 3.2129 - 3.2046 |
| Cyclic AMP | HMDB0000058 | 8 | 1 | 6.1548 - 6.1498 |

| | | | | |
|---|---|---|---|---|
| D-Glucose | HMDB0000122 | 62 | 1 | 3.8952 - 3.8917 |
| D-Glutamic Acid | HMDB0003339 | 36 | 2 | 2.343 - 2.3384 |
| Dimethylamine | HMDB0000087 | 1 | 1 | 2.7313 - 2.735 |
| D-Mannose | HMDB0000169 | 10 | 2 | 3.3712 - 3.3685 |
| Ethanol | HMDB0000108 | 3 | 2 | 1.1995 - 1.195 |
| Formic acid | HMDB0000142 | 1 | 2 | 8.4626 - 8.4562 |
| Glutathione | HMDB0000125 | 13 | 2 | 2.5718 - 2.5614 |
| Glycerol | HMDB0000131 | 7 | 2 | 3.6449 - 3.6424 |
| Glycol | HMDB0001881 | 2 | 2 | 1.1443 - 1.1381 |
| GTP | HMDB0001273 | 3 | 2 | 5.9546 - 5.9465 |
| Indoleactic acid | HMDB0000671 | 5 | 2 | 7.7515 - 7.7414 |
| Inosinic acid | HMDB0000175 | 5 | 2 | 6.1629 - 6.1548 |
| Isobutyric acid | HMDB0001873 | 1 | 2 | 1.1137 - 1.1082 |
| Isopropyl alcohol | HMDB0000863 | 3 | 2 | 1.1847 - 1.1759 |
| Lactic acid | HMDB0000190 | 6 | 1 | 4.1209 - 4.1167 |
| L-Alanine | HMDB0000161 | 5 | 1 | 1.4826 - 1.4768 |
| L-Arginine | HMDB0000517 | 28 | 2 | 1.6403 - 1.6349 |
| L-Aspartic.acid | HMDB0000168 | 17 | 1 | 2.8595 - 2.8542 |
| L-Glutamine | HMDB0000641 | 26 | 1 | 2.4785 - 2.4733 |
| L-Histidine | HMDB0000177 | 2 | 1 | 7.0845 - 7.0758 |
| L-Isoleucine | HMDB0000172 | 21 | 1 | 1.0154 - 1.0059 |

| | | | | |
|---|---|---|---|---|
| L-Leucine | HMDB0000687 | 28 | 1 | 0.9695 - 0.9613 |
| L-Lysine | HMDB0000182 | 28 | 2 | 3.0323 - 3.0275 |
| L-Methionine | HMDB0000696 | 12 | 1 | 2.6355 - 2.6264 |
| L-Phenylalanine | HMDB0000159 | 11 | 1 | 7.3427 - 7.3364 |
| L-Proline | HMDB0000162 | 12 | 2 | 3.3052 - 3.3001 |
| L-Serine | HMDB0000187 | 23 | 2 | 3.9558 - 3.9528 |
| L-Threonine | HMDB0000167 | 8 | 1 | 4.2574 - 4.2538 |
| L-Tyrosine | HMDB0000158 | 9 | 2 | 6.9145 - 6.9107 |
| L-Valine | HMDB0000883 | 13 | 1 | 0.9934 - 0.9867 |
| Methyl Acetate | HMDB0015502 | 1 | 2 | 1.9225 - 1.9169 |
| Myo-Inositol | HMDB0000211 | 14 | 1 | 3.2853 - 3.2821 |
| NAD+ | HMDB0000902 | 10 | 1 | 9.1575 - 9.1485 |
| NADH | HMDB0001487 | 18 | 2 | 8.2099 - 8.2052 |
| NADP+ | HMDB0000217 | 10 | 2 | 8.1921 - 8.184 |
| Phosphocholine | HMDB0001565 | 1 | 2 | 3.2284 - 3.2211 |
| Propionic acid | HMDB0000237 | 9 | 2 | 1.0745 - 1.0659 |
| Pyroglutamic acid | HMDB0000267 | 30 | 2 | 2.5178 - 2.5141 |
| Saccharopine | HMDB0000279 | 46 | 2 | 3.0819 - 3.0742 |
| Sarcosine | HMDB0000271 | 2 | 2 | 2.234 - 2.239 |
| Taurine | HMDB0000251 | 9 | 1 | 3.4164 - 3.4124 |

**Supplementary Table 2. PLS-DA validation table.** Percentage score of all assessed metrics in the machine learning model with their standard deviation. UNTR = untreated, JAKi = pan JAK inhibitor I, BARI = Baricitinib, TOFA = Tofacitinib.

| | **Accuracy** | **Balanced Accuracy** | **F1 Score** | **Precision** | **Recall** |
|---|---|---|---|---|---|
| UNTR 0h | 90% (13.7) | 90% (13.7) | 86.7% (18.3) | 100% (0) | 80% (27.4) |
| UNTR 2h | 95% (11.2) | 95% (11.2) | 93.3% (14.9) | 100% (0) | 90% (22.4) |
| JAKi | 85% (13.7) | 85% (13.7) | 80% (18.3) | 100% (0) | 70% (27.4) |
| BARI | 95% (11.2) | 95% (11.2) | 93.3% (14.9) | 100% (0) | 90% (22.4) |
| TOFA | 95% (11.2) | 95% (11.2) | 93.3% (14.9) | 100% (0) | 90% (22.4) |

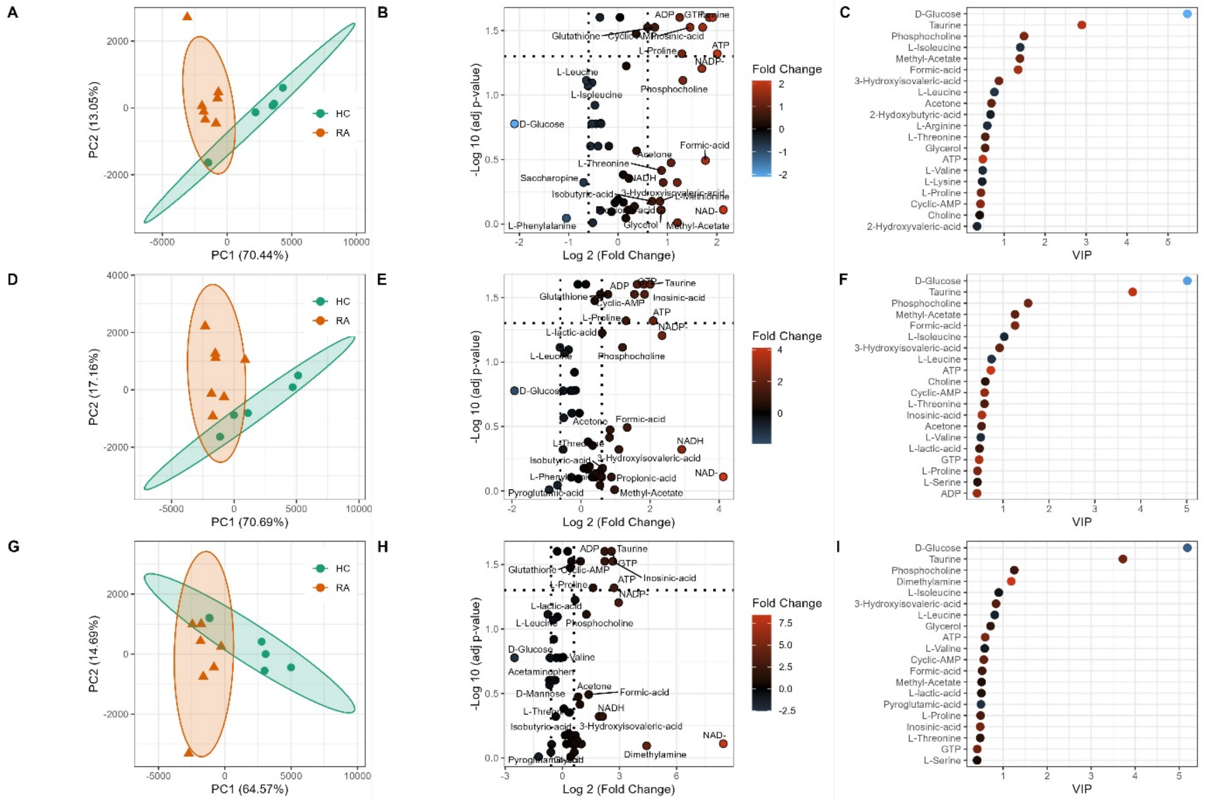


**Supplementary Figure 1 Metabolomics differences between RA and HC neutrophils with JAK inhibitors**. PCA scores plot of HC and RA neutrophils with pan-JAKi (A) Baricitinib (D) and Tofacitinib (G) showing separation on the first principal component (PC). Volcano plot (B,E,H) showing metabolites significantly different between HC and RA neutrophils (adj p-value < 0.05) and the log2 fold change (FC) for each metabolite as indicated by gradient colour scale provided. Variable importance in projection (VIP) (C,F,I) obtained from PLS-DA showing top 20 metabolites for each model.

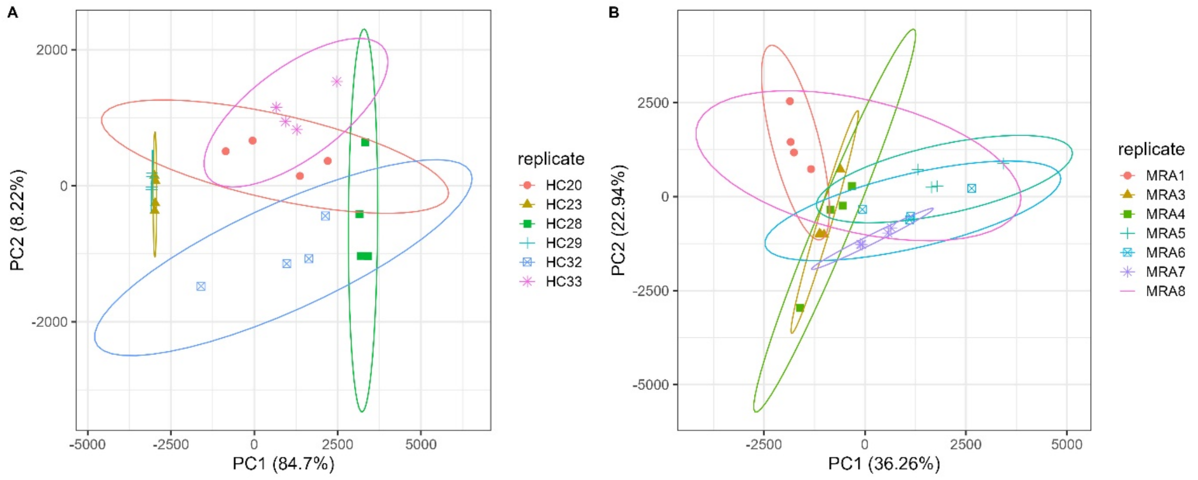


**Supplementary Figure 2. PCA showing high between-subject variability of neutrophils treated with JAK inhibitors.** (HC = healthy control, RA = rheumatoid arthritis).

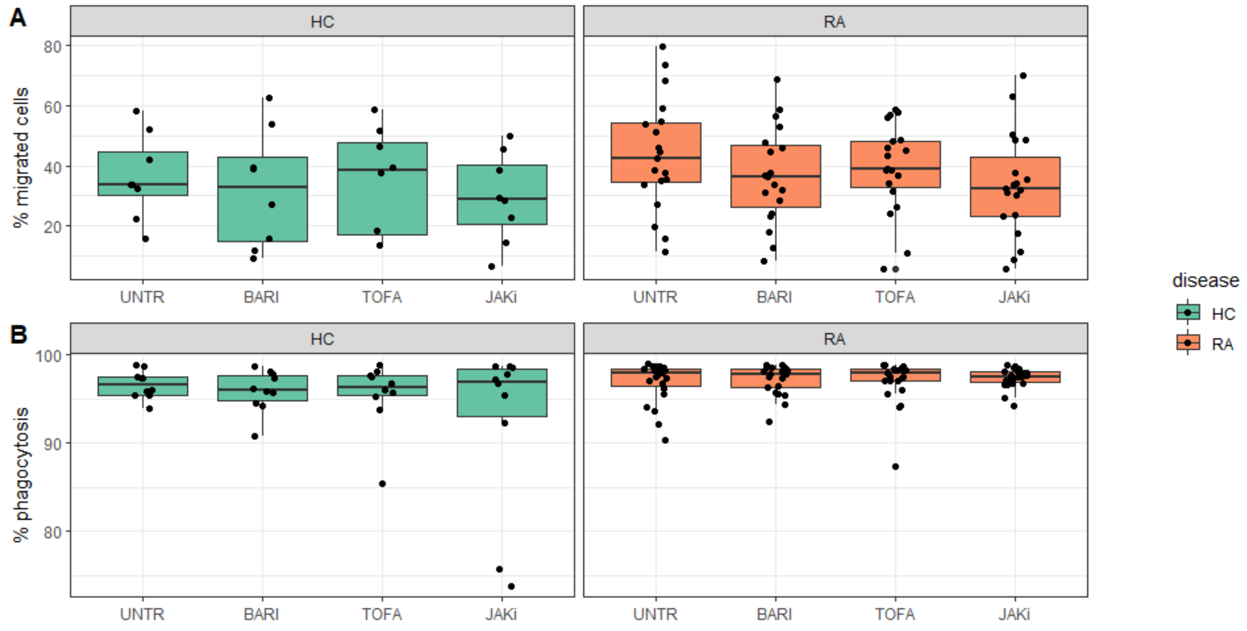


**Supplementary Figure 3. Effect of JAK inhibitors on neutrophil migration and phagocytosis.** (A) Effect of JAK inhibitors on neutrophil chemotaxis towards fMLP. Neutrophils were pre-treated with JAKi for 30 min and then allowed to migrate towards fMLP ($10^{-8}$M) for 90 min. (B) Effect of JAK inhibitors on neutrophil phagocytosis. Neutrophils were pre-treated with JAKi for 30 min and then incubated with FITC-labelled latex beads for 30 min. None of the inhibitors tested prevented phagocytosis or cell migration. UNTR = untreated, JAKi = pan JAK inhibitor I, BARI = Baricitinib, TOFA = Tofacitinib.
